# Perspectives of HIV policy makers and program implementers regarding the design and utilization of HIV surveillance systems in Sub-Saharan African countries experiencing a declining HIV epidemic: a qualitative study

**DOI:** 10.64898/2026.02.05.26345633

**Authors:** Victor Mwapasa, Fumbani Chigawa, Chikondi Mafaiti, Marriott Nliwasa, Takondwa Charles Msosa, Jeffrey W. Imai-Eaton, Beth A Tippett Barr

## Abstract

**Background:** The burden of new HIV infections and HIV-related deaths have declined dramatically in sub-Saharan Africa (SSA). However, current HIV surveillance systems are primarily donor-funded and rely on data from population-based surveys and routine health services. These need to evolve so that they can reliably monitor future HIV trajectory, in the context of declining burden of HIV and limited donor funding. We qualitatively assessed stakeholder perceptions of the current status and future needs for HIV surveillance.

**Methods:** From September 2024 to February 2025, we conducted a grounded-theory qualitative study whose participants were representatives of international HIV agencies, policy makers and health sector development partners and HIV programme managers from Malawi, Lesotho, Zimbabwe, Ghana and Kenya, and HIV programme implementers from Malawi. We conducted 28 online and in-person key informant interviews and three focus group discussions with 34 Malawi-based participants working at sub-national level. These were audio-recorded for transcription. We conducted sequential deductive and inductive content analyses.

**Results:** We found that HIV programs in SSA are familiar with and have successfully used routine health system data, population-based surveys, and mathematical modeling for HIV surveillance and monitoring and evaluation (M&E). However, most respondents could not distinguish the differences between M&E and surveillance and were unaware of the key inputs for mathematical models used to estimate key impact indicators. They expressed concern over the parallel HIV data systems, lack of integration with the broader health surveillance systems, sub-optimal quality of routine facility-based data, and the huge cost and limited precision of population-based surveys. They recommended investment in several areas including data quality improvement, adoption of digital technology and artificial intelligence to improve the efficiency of the surveillance system, expanded stakeholder sensitization in mathematical modeling, implementation of targeted surveys focusing on high-risk populations, and prioritization of HIV morbidity and mortality indicators.

**Conclusions:** Future HIV surveillance strategies need to invest in institutionalizing local capacity for using multiple HIV data streams to inform key surveillance indicators through modeling and analytic tools, establishing management systems to enhance routine data quality, streamlining HIV surveillance and M&E indicators, and fostering disease surveillance integration. Targeted surveys will be required to complement routine facility-based surveillance.

## Introduction

UNAIDS estimates that between 2010 to 2024 the number of new HIV infections declined by 40% and AIDS-related deaths by 54% globally [1]. In sub-Saharan Africa (SSA), home to the majority (60%) of people living with HIV (PLHIV), progress at reducing infections and deaths have been even greater at 56% and 59%, respectively [2]. By 2024, in Southern and East African countries, 93% of PLHIV were aware of their status, 84% were on lifelong ART, and 80% of all PLHIV were virally suppressed, with seven countries already achieving the UNAIDS 95-95-95 targets [2]. Nevertheless, over the past 5 years, the rate of decline of HIV infections and AIDS-related deaths have slowed in most SSA countries, slower than what is required to achieve global target of 90% reduction by 2030, and sustain HIV epidemic control beyond 2030. Robust HIV surveillance systems will be required to effectively track small changes in national and sub-national trends and distribution of HIV infections and deaths and guide timely adjustments to programmes and intervention strategies to prioritize the geographical areas and populations with unmet prevention and treatment needs.

In the 1980’s, HIV surveillance in SSA relied on AIDS case reports, followed by establishment in the 1990s of sentinel surveillance of HIV prevalence among pregnant women attending antenatal clinics, along with other populations at higher risk, including sexually transmitted infections (STI) clinic attendees and sex workers [3]. More recently, surveillance has mostly relied on population-based surveys and data from routine health system service delivery, interpreted through mathematical models such as Spectrum and Naomi [4, 5]. While routine health facility data provide information on HIV service utilization, by default it excludes sub-populations who are less likely to access health facilities, and who may bear a disproportionate HIV burden. In contrast, population-based surveys such as Demographic and Health Surveys (DHSs) and Population HIV Impact Assessment Survey (PHIAs) [6, 7] provide representative data on HIV burden and risk factors in the general population, and the Integrated Biological and Behavioural Surveillance Surveys (IBBSs) focus on high-risk sub-groups. However, these surveys are conducted every 3-5 years, and they do not provide the frequency or timeliness of results needed to facilitate programmatic decision-making. They are also expensive and almost exclusively donor-funded, and therefore may not be sustainable in future, particularly in the current context of declining development assistance.

Mathematical models which triangulate data from routinely collected health programme data and population-based surveys provide up-to-date estimates of the burden of HIV and coverage of treatment interventions, thereby facilitating programmatic decision-making. However, the reliability and accuracy of these models depend on the quality of routine health service data and continued availability of population-based data.

As the HIV epidemiological context and funding landscape continue to change, national HIV Monitoring and Evaluation (M&E) frameworks and HIV Surveillance Strategies in SSA may need to evolve accordingly. In view of the ongoing and anticipated changes in HIV epidemiology, service delivery and data collection approaches, countries need to design HIV surveillance systems that sustainably produce reliable, accurate and timely HIV estimates at national and sub-national levels. As a foundation for designing future HIV surveillance strategies with this level of rigour, which can monitor sustained HIV epidemic control in SSA, we conducted a study with the objectives to:

1. Describe HIV surveillance stakeholders’ views on the appropriateness and adequacy of current HIV surveillance strategies in tracking HIV epidemic changes
2. Explore stakeholders’ perspectives on the current utilization of HIV surveillance findings for programmatic decision-making, and
3. Describe surveillance experts’ views on how HIV surveillance methods and indicators should change in response to an evolving HIV epidemic.

## Methods

### Study Design and Population

From September 2024 to February 2025, we conducted a grounded-theory qualitative study involving HIV programme stakeholders working in Malawi, Zimbabwe, Ghana, Kenya, Lesotho, and international HIV funding and technical support agencies. At global level, we approached HIV policy and technical experts from national HIV programs and institutes such as the World Health Organization (WHO), the Joint United Nations Programme on HIV/AIDS (UNAIDS), Africa Centers for Disease Control (Africa-CDC), the United States Centers for Disease Control and prevention (US-CDC), and the President’s Emergency Plan for AIDS Relief (PEPFAR). At national level in the five countries, participants included policy makers, development partners, programme implementers, M&E managers, representatives of civil society organizations, and technical advisors. In Malawi, we also included implementers of HIV and other health programs at sub-national level. Eligible participants were at least 18 years old and had more than one year of experience working in HIV programmes in the public, private and NGO sectors, or in bilateral and multilateral agencies.

### Study Setting

The study was conducted in Lilongwe, the capital city of Malawi, where major policy makers and development partners are based and in purposively selected high HIV burden districts of Blantyre, Thyolo and Mangochi in Southern Malawi, where major HIV implementing partners are based. Virtual interviews were conducted with regional and global experts.

### Sample Size and Sample Selection Procedures

We planned to conduct key informant interviews (KIIs) with up to 21 participants from Malawi and 19 from the four SSA countries and global stakeholders. In Malawi, we first identified potential respondents from the HIV Monitoring, Evaluation and Surveillance Technical Working Group (MERS TWG), which consists of representatives from the Ministry of Health, development partners, civil society, and HIV programme implementation partners. We identified additional respondents based on recommendations from TWG members, until the targeted number and composition of KIIs was achieved. We also conducted three focus group discussions (FGDs), one in each of the three high HIV burden districts, with individuals responsible for implementing and supporting the delivery of HIV programmes. Each FGD consisted of a minimum of 10 participants.

### Data collection procedures

Before conducting KIIs and FGDs, the study team conducted a desk review of existing HIV surveillance strategies, guidelines, systems and indicators used international, regionally and globally [8]. Through this review, the team identified HIV surveillance methodologies, indicators and data collection platforms currently used for tracking the trajectory of HIV in different settings. Findings from the desk review were used to develop themes for further interrogation in this qualitative study. For example, the qualitative study focused on the stakeholders’ experiences in implementing HIV surveillance guidelines and methodologies and the utilization of surveillance data for decision-making.

Data collectors included the study coordinator, co-investigators and trained master’s students who co-developed the study protocol. Before commencement of data collection, they were trained in research ethics and the implementation of study tools. We also sensitized leaders of key stakeholder institutions from whom our study participants were drawn, through the MERS TWG. Interview guides were used to conduct the KIIs and FGDs. These included broad themes such as awareness and utilization of surveillance reference documents, strengths and weaknesses of surveillance systems and data sources, ease or complexity of generating HIV surveillance indicators, level of utilization of surveillance data and future priority HIV surveillance indicators, systems and data sources. Each of these thematic areas included appropriate probes (Supplementary material #1). Data collection was conducted either face-to-face or virtually for KIIs, and face-to-face for FGDs. Efforts were made to conduct the interviews/discussion in a private place away from distractions. All interviews and discussion were conducted in English and were audio-recorded. In line with our institution’s ethical requirements, study participants received appropriate monetary compensation for their time, efforts and disruption from normal day-to-day activities to participate in the study and aligned with their own organizational policies.

### Data management and analysis

Audio-recorded interviews were transcribed using an appropriate and approved artificial intelligence (AI)-based transcription service, Sonix^TM^ (https://sonix.ai/research-firms). All electronic files were anonymized. Participants were identified only by a unique participant ID number. Each transcript was reviewed, checked for quality and cleaned by trained research assistants supported by the study co-investigators. Data were stored on a secure SharePoint server with access restricted to study co-investigators. All documents were stored securely and only accessible by study staff and authorised personnel.

Thematic content analysis of Microsoft Word transcripts was conducted using NVivo (QSR International Pty Ltd, Australia) software. Initially, deductive content analysis was conducted using the themes already identified in the interview guides. Thereafter, NVivo was used to conduct inductive content analysis by identifying codes, or repeated words and phrases used by participants, and then grouping them into themes and sub-themes. The lead investigators reviewed the findings to ensure consensus was reached on the meaning and relative importance of each code and theme.

### Ethical Considerations

The protocol, informed consent form, and participant information sheet were reviewed and approved by the College of Medicine Research Ethics Committee (Approval No: P.04/24-0665). Before the interviews, informed consent was obtained from KII participants by written or email consent. Copies of the informed consent form describing the purpose of the study, data collection process and how their rights are safeguarded, were shared with the study participants. Participants were advised that they were free to withdraw from the study at any time, with no reason required, and with no negative consequences.

## Results

### Study participant characteristics

From 10^th^ September 2024 to 4^th^ February 2025, we approached 21 national and 19 international respondents for KIIs. Of these 16 (76%) national and 12 (63%) international respondents consented and were interviewed. Of the 12 international participants, eight were from Kenya, Zimbabwe, Ghana and Lesotho and four were from international bilateral and multilateral development partners. We approached 36 respondents at district level to participate in FDGs, of whom 34 (94%) consented. Most KII participants were M&E and surveillance experts and had postgraduate degrees (Table 1). The FDG participants were primarily HIV coordinators and service providers, and the majority had bachelor’s degrees or diplomas.

**Table 1.**
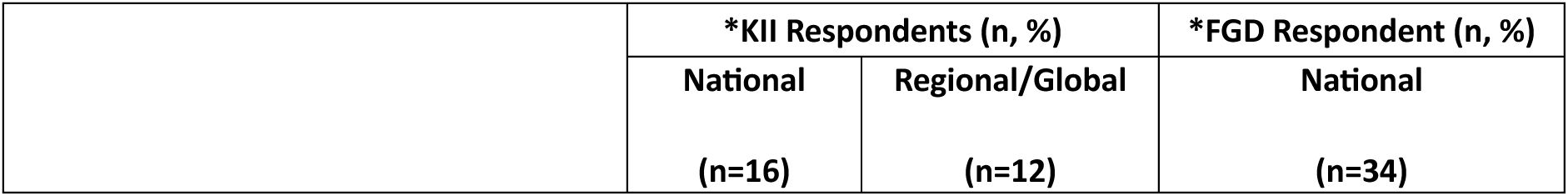
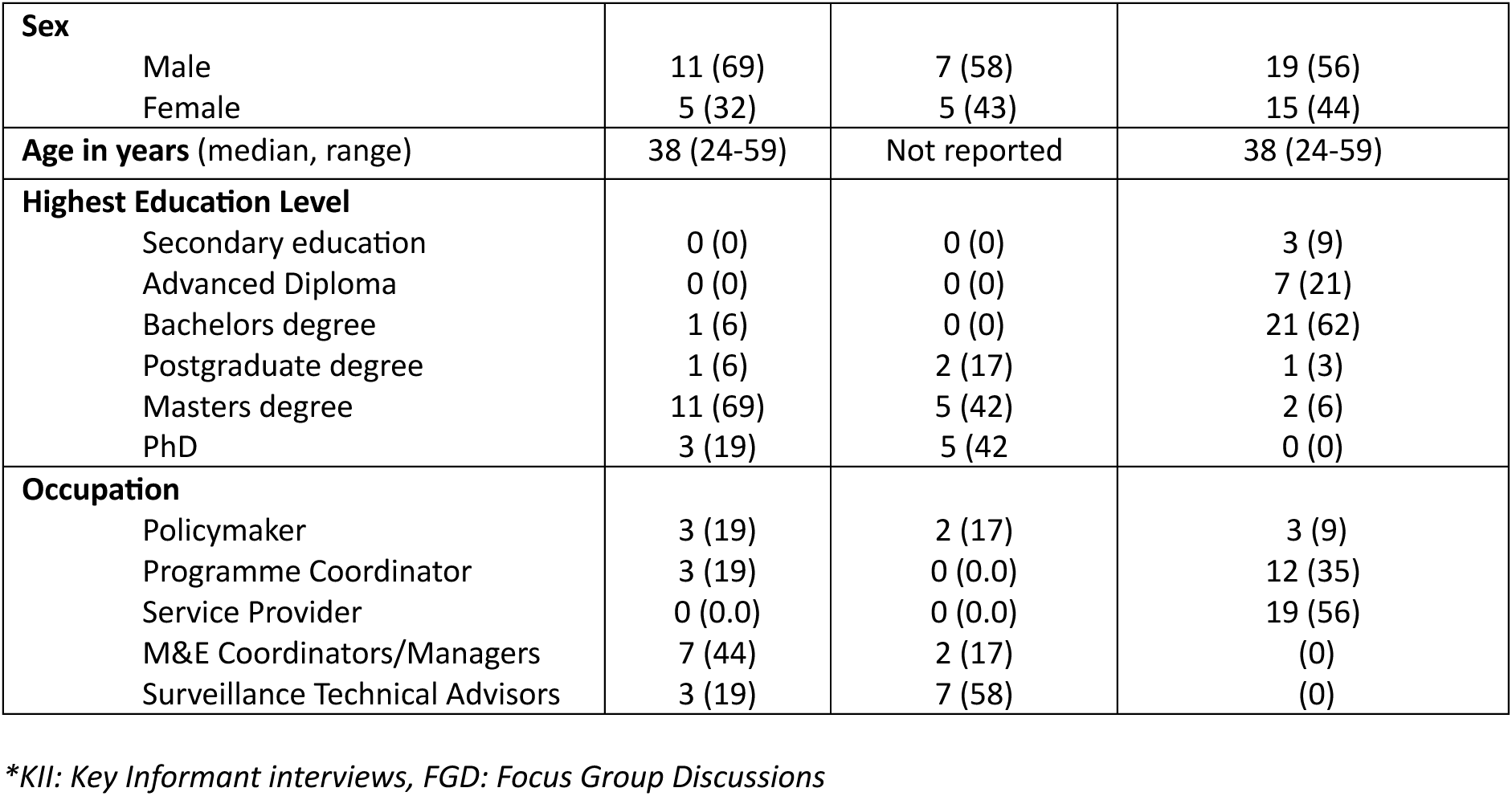
Respondent characteristics.

Several themes emerged within each study objective (Table 2), with similarities in data use and reported data needs by participants engaged at national and sub-national levels, and a deeper technical understanding of surveillance and disease modelling among participants working at regional level and in multilateral agencies than those at national and sub-national level.

**Table 2.** Emerging themes with the study objectives.

| Study Objective | Theme |
| --- | --- |
| 1. Stakeholder perspectives on current HIV surveillance methods | 1.1 Multiple, poorly harmonized HIV surveillance guidelines and data sources |
|  | 1.2 Suboptimal integration between HIV and other disease surveillance. |
|  | 1.3 Few strengths and many weaknesses of current HIV surveillance methods |
| 2. Stakeholder utilization of HIV surveillance data for decision-making | 2.1 Uncertainty around the difference between surveillance and M&E |
|  | 2.2 Lack of awareness around data needs for modeling |
|  | 2.3 Predominant use of data for M&E and not surveillance |
| 3. Priorities for future HIV surveillance methods and indicators | 3.1 Strengthening facility-based surveillance |
|  | 3.2 Tailored surveillance approaches |
|  | 3.3 Leveraging individual-level electronic data system |
|  | 3.4 Conflicting views on the importance of mortality surveillance |
|  | 3.5 Streamlined HIV surveillance indicators |

### Objective #1: Stakeholder perspectives on the appropriateness of current HIV surveillance methods

Overall, respondents offered wide-ranging views regarding the meaning of “surveillance” and on the appropriateness, adequacy, importance and challenges of their current HIV surveillance systems. These were grouped into three key themes:

#### 1.1 Multiple parallel surveillance reference documents and reporting systems

National KII respondents reported using multiple reference documents for HIV surveillance, including the National Strategic Plan for HIV and AIDS (NSP), Health Sector Strategic Plan (HSSP) and Ministry of Health (MoH) integrated guidelines for HIV testing, ART and PMTCT. In addition, respondents from non-governmental implementing partners mentioned donor-specific reference documents such the USAID Indicator Reference Manual. At regional and global levels, respondents cited the UNAIDS, WHO and PEPFAR guidelines and the Global AIDS Strategy as key surveillance reference documents. Respondents tended to reference the surveillance systems that were relevant to their field of work. For example, Population-based HIV Impact Assessment surveys (PHIA) were mentioned by national program planning officers, Biological and Behavioral Surveillance Surveys (BBSS) were mentioned by those in key populations work, and HTS and ART guidelines by those working in the HIV testing and treatment programs.

In addition to the reference documents, participants described several data sources and platforms for HIV surveillance, including the Demographic and Health Survey (DHS) and PHIA surveys, electronic medical records (EMR), the Department of HIV Management Information System (DHAMIS) and the District Health Information System (DHIS-2). National and regional respondents also expressed frustration with the existence of multiple donor-funded HIV surveillance studies, data collection systems, and reporting obligations. The parallel systems were reported to overburden health workers and to produce conflicting statistics.

> *“Each funder seems to have its own platform. You have to understand their indicators, their frequency of reporting, and their verification processes. It’s hard to align with the national priorities when you’re always chasing donor-specific requirements.”* —District M&E Officer, Malawi

> *“There are so many systems running in parallel. There’s DHIS2, there’s DATIM, there’s the EMR, and sometimes you’ll find discrepancies in numbers because they are pulling from different timeframes or not cleaned properly. It makes it hard to know what to trust.”* —National-level M&E Officer, Malawi

> *“We report to NAC using one format, then to PEPFAR using DATIM, and to the Global Fund using another tool altogether. It becomes difficult for us to manage the reporting burden, especially with limited staff.”* —Implementing partner, Malawi

> *“We have a problem of parallel systems in Lesotho. You know, the implementing partners have to report to the donors. And because the systems are not very stable… they tend to use their own systems so that they’re able to meet their reporting requirements. This hinders the focus from ensuring that the national system actually is fully supported.”* —Policymaker, Lesotho

#### 1.2 Sub-optimal integration between HIV and other disease surveillance

No respondents mentioned the existence of a standalone HIV surveillance strategy or reported ever using existing integrated government surveillance systems or platforms, such as the Integrated Disease Surveillance and Response system or One Health Surveillance Platform, for reporting HIV-related data. A few respondents noted the health system’s over-reliance on donor-funded M&E systems and the underutilization of the national DHIS-2 system for HIV reporting despite having important features that allow individual level reporting (e-Tracker) and ordering of pharmaceutical and medical supplies. In their view, this represented a lost opportunity for strengthening country-owned integrated reporting and procurement systems. However, some FGD participants from one of the districts expressed satisfaction with using a bespoke integrated electronic data analytic system called PALMS (Preventive Adaptive Learning Management Systems) that provides monthly visual dashboards of HIV and other health indicators at primary care health level, facilitating timely identification of poor indicators.

#### 1.3 Few strengths and many weaknesses of current HIV surveillance methods

Most KII respondents acknowledged that HIV surveillance is critical for informing HIV program decision-making. Key advantages of existing systems included easy interpretation of surveillance data to guide decision-making, the generalizability and reliability of findings from population-based studies, and their ability to identify new infections and high-risk populations. However, the majority of respondents, within and outside Malawi, expressed concern over the suboptimal quality of routinely collected health facility data, characterized by duplicate entries, poor and inconsistent data cleaning at the point of entry, untimeliness of data from population-based surveys, and misalignment between donor data collection and reporting systems and the government’s integrated M&E system (Table 3). Several respondents noted the unavailability of variables for capturing data on key and priority populations such as men who have sex with men (MSM), female sex workers (FSW), prisoners, fisherfolk, young people, and people with disabilities.

**Table 3:** Perceived weaknesses of current HIV surveillance methods.

| Disadvantages | Illustrative quotes |
| --- | --- |
| **Facility-based surveillance** |  |
| Delayed and inaccurate data for decision-making | *"The current measurements of viral load ..... we do not have systems that would enable us to have viral [load] results with the shortest turnover [sic] time. So... results will come about 3 or 4 months later. We are bound to miss certain things."* —National implementing partner, Malawi |
| Sub-optimal quality characterized by data incompleteness, deliberate over-reporting, inability to uniquely identify, inadvertent double or triple counting and an inability to disaggregate old versus new infections | *"... But this data is coming from paper-based systems which naturally have got challenges in terms of accuracy of using health facility data in national picture, because they are not very sensitive to issues like transfers, issues like duplicates.. —National program donor, Zimbabwe*<br>*"Some patients, quite wisely, I think, register themselves at multiple clinics in order to avoid stockouts. So then they show up as multiple patients when they're not really — they're really one patient." —Global expert*<br>*".....because of [implementing partners] chasing the numbers ..... to ensure that they achieve the targets.... sometimes they compromise the quality of the services." —National Policymaker, Malawi* |
| Cannot be generalized | *"Our data source has been routine patient registers. That's the data source that we've been relying on. But that data is biased. We are not able to access everyone's data simply because some of them don't have access to health facilities" —National implementing partner, Malawi* |
| Limited sub-population disaggregation | *"Within the routine system I may not know surely, but it will be really hard to determine whether that somebody is a key population or not and where they are identified. The challenge will be now getting a sufficient data for your estimation, since these are rare populations." —National implementing partner, Malawi* |
| **Population-based surveys** |  |
| Unsustainably expensive and donor-dependent | *"PHIA helped us reorient the program, but it's very expensive. Now with EMRs and unique IDs, we're questioning if we still need them." —National donor, Kenya* |
| Survey infrequency and untimeliness of findings | *"The challenge with the IBBS surveys in the past is that they only occur every, I don't know, 3 to 5 years. And then the results come late, almost by the time they're analyzed are sort of out of date." —Global expert* |
| Missing data on key and priority populations | *"IBBS helps us reach the known key populations, but it doesn't help us identify new or missed groups like fisherfolk or truckers." - Global expert*<br>*"One of the national household surveys used to be DHS. .... The limitations there are, you know, it only happens every five years. ... Children are not included. So we have some big question marks around our estimates of number of HIV positive children." —Global expert* |
| Can not generate precise estimates when the prevalence is low | *"...the prevalence has gone down. The precision of most of these surveillance activities is becoming questionable and it is more expensive to have precise estimates from population-based surveys.... over time when you look at the results, then they are not as robust as we used to have before... it's about time we start thinking through to say, how best do we carry out these kinds of surveys for the future." —National program donor, Kenya* |
| **Mathematical Models** |  |
| Inaccuracy due to poor quality data | *"We think it's fairly decent, but it is a modelled estimate... And as I mentioned, as we have some questions about the quality of the ART program reports... any discrepancies in their program data will be quite apparent. In Zimbabwe... they have a few provinces that apparently have more than 100% coverage of ART. Well, something is wrong somewhere." —Global Expert* |
| Inaccurate population denominators which likely results in errors when | *"..... I think we've done a very good job in terms of trying to account for the numbers on treatment. But at the denominator level, I think to me the challenge would be how will the estimates team relook at that process and* |
| estimating HIV incidence and prevalence. | *use all existing data to see how best to refine [denominator estimates]" —*<br>National program donor, Kenya |

Respondents frequently mentioned mathematical models, such as Spectrum and Naomi, as a key surveillance data sources for estimating HIV infections and deaths, although respondents were not commonly familiar with the surveillance and M&E data inputs required for the models, or how key outputs from the models relate to specific input data. One regional respondent questioned the accuracy of the Spectrum model in estimating the burden of HIV infections in children and the number of HIV-related deaths.

Several respondents noted efforts made by their countries to implement mortality surveillance, which could potentially generate good data on AIDS-related mortality. However, most expressed concern that mortality surveillance was poorly implemented at both health facility and community levels. Whilst the basic framework and system for mortality surveillance existed in most countries, the respondents noted that the system was generally poorly resourced and was characterized by poor reporting and lack of oversight.

> *So in Zimbabwe…in each and every district … there is an office of civil registration which registers all births and deaths…a person in Zimbabwe cannot be buried without a burial order. …So we have got the community registration of all community deaths and also of facility. However, we tend to have incomplete records of our community deaths in terms of number one, the cause of death.* —National program donor, Zimbabwe

### Objective #2: Stakeholder utilization of HIV surveillance data for decision-making

There was broad concurrence on the usefulness of HIV data to guide programmatic decision-making for both national HIV programs and implementing partners. However, various observations were made around the following sub-themes:

#### 2.1 Uncertainty around the difference between M&E and surveillance

At national and sub-national levels, many respondents did not clearly distinguish between monitoring and evaluation (M&E) and surveillance methods or purposes. Respondents frequently referred to using HIV data to monitor the immediate outputs of their specific HIV interventions (M&E), and not to assess changes in the trajectory and distribution of the epidemic (surveillance). For example, although some local respondents reported using antiretroviral therapy (ART) electronic medical records (EMR) data for surveillance, only a few reported analysing the data to identify the sub-populations with a high burden of HIV infections or at increased risk of mortality. Most stakeholders used the EMR primarily to quantify the number of ART initiations and to identify treatment defaulters and the number of clients retained in care. Statements by a few revealed their knowledge of more precise difference between M&E and surveillance:

> *” the data from EMR hasn’t been used for surveillance yet. But what we have been using it is for mainly reporting and program improvement purposes.”* —National program donor, Malawi

> *“….there’s still confusion in some quarters between surveillance and M&E—many people think of surveillance only as what comes through routine reporting, not understanding the broader elements like surveys or modelling.”* —National program donor, Ghana

#### 2.2 Lack of awareness around data needs for modeling

Most respondents acknowledged the importance of annual mathematical models in estimating the number of new infections and HIV-related deaths and estimating program ART coverage. However, various statements from the respondents revealed limited participation outside of MOH and NAC in producing annual estimates. Few respondents at both local and international levels, could clearly identify or explain the key data points or their sources which are required to run and calibrate the models. Most respondents simply mentioned the “HIV Estimates Group” as being responsible for running the models and collating necessary input data. There was limited understanding that mathematical models combined routine program data with population-based survey data to model annual incidence and mortality.

> *“…..I’m aware of what they call Spectrum. Yes, I have used Spectrum before because there was some information that I wanted… but I’m not sure who else is part of it.”* —National Program Officer, Malawi

> *“Apart from the MPHIA, I think we have what we call HIV spectrum. I think it’s done by DHA and NAC? I’m not sure who else is part of it, but it’s shared regularly, and we normally use that to see the current estimates and how we fit in.”* —National policymaker, Malawi

> *“I think they produce a report from the Department of HIV. I think it’s the Spectrum. It’s produced annually. Yeah. I think that would be a very good data source. I feel like it’s comprehensive enough.”* —National policymaker, Malawi

#### 2.3 Predominant use of data for M&E and not surveillance

Most stakeholders acknowledged the critical role that a robust surveillance system plays in the implementation of HIV programs. They reported numerous ways in which surveillance data are utilized (Table 3), however, it was evident that surveillance system outputs were primarily used for M&E and, to a lesser extent, for HIV surveillance. Nevertheless, it was noted that the Recency HIV surveillance program, piloted by the US CDC in selected health facilities, is being used not only for identifying recently infected individuals but also mapping HIV transmission hotspots and facilitating targeted interventions.

**Table 3:**
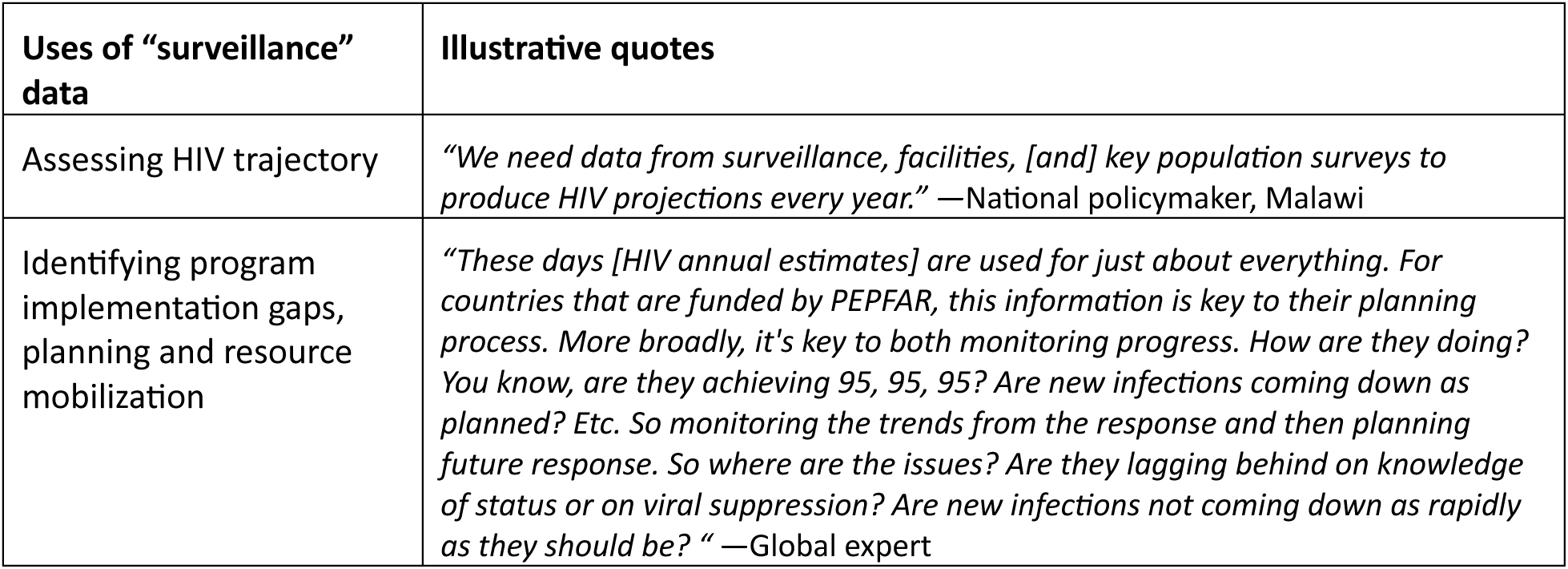
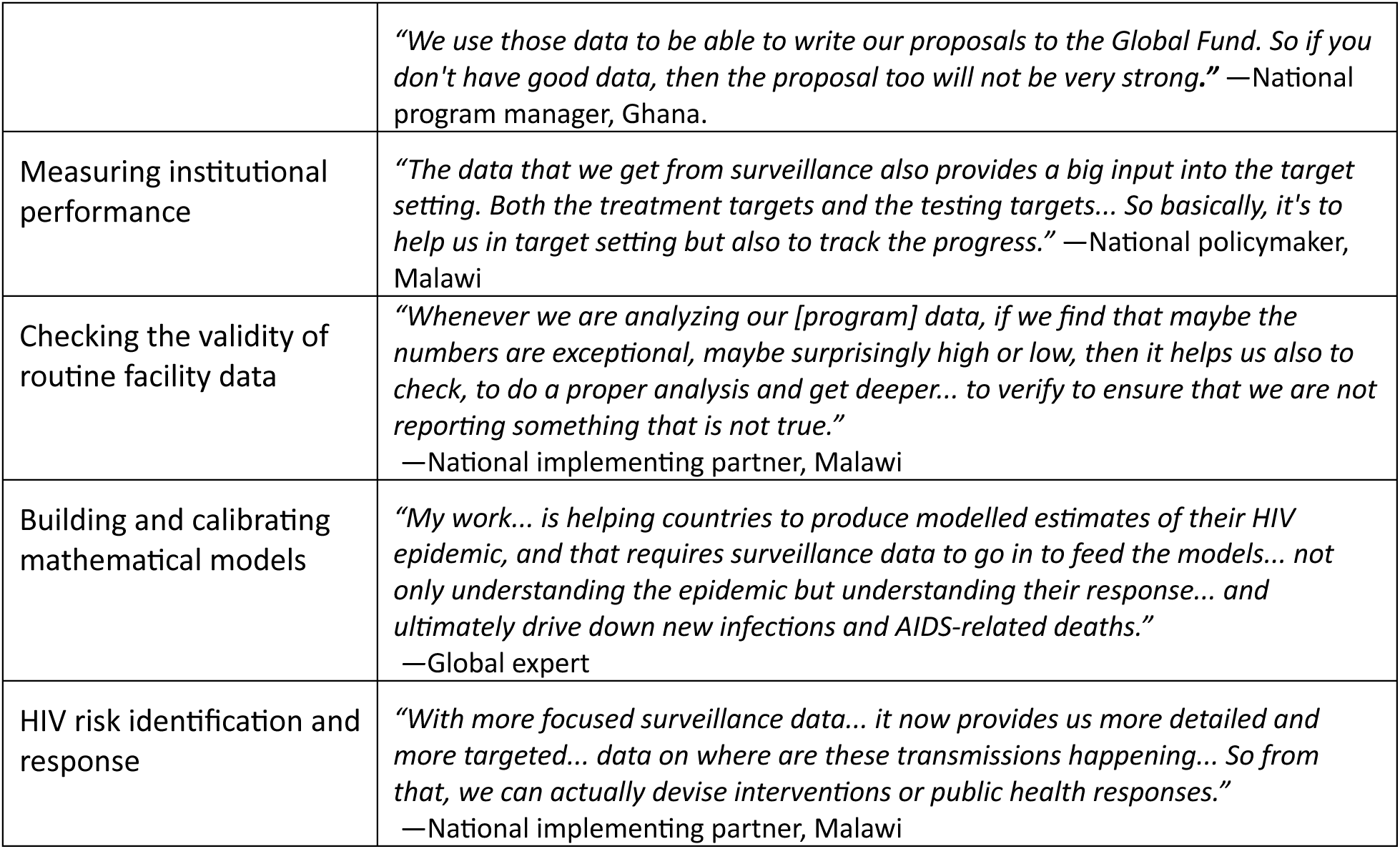
Reported uses of “surveillance” data.

### Objective #3: Priorities for future HIV surveillance methods and indicators

Most respondents acknowledged that the current HIV surveillance system, despite its weaknesses, had been effective in monitoring the trajectory of HIV and facilitating targeted responses and were of the view that no changes were required. They were generally unsure whether the surveillance methods and systems are appropriate or adequate to detect smaller incremental changes in key HIV impact indicators that are currently observed and may continue in future. Nevertheless, various regional and international respondents made various suggestions for improving the surveillance system in future:

#### 3.1 Need to strengthen facility-based surveillance

Many respondents believed that routine, high quality health facility surveillance will remain the backbone of any future HIV surveillance system, and therefore investment is needed in several strategic areas, including human resources, infrastructure, data quality assurance, unique identifier numbers for clients, digital and artificial intelligence, and integration of HIV within other disease surveillance programs.

> *“We have to recognize that going into the future, I think it’s really going to be important to continue to improve the routine data systems. And I think that’s what we’re going to be relying on in the future. […] So I think if we can get the routine testing, the routine program data in good shape, […] that will go a long way towards what we need in the future.“* —Global expert

> *“UNAIDS has been encouraging countries to do data quality assessments. Countries that have done that have usually ended up reducing their estimates of number of people living with HIV or on ART by ten, fifteen percent or so.”* —Global expert

> *“The country is trying to pursue a health number to use it as a unique ID so that we can move away from what currently we have got unique numbers in terms of different programs. So we are trying to generate […] the National Health ID number […] but the progress has been very slow […] and I have not seen its utility to date. […] But the equipment is not even yet there.”* —National program donor, Zimbabwe

> *“We have a problem of parallel systems in Lesotho […] demolishing the parallel system [is essential] so that we focus on one national M&E system. Otherwise, we believe Lesotho is on the right track. We just have to ensure that we work on the challenges that we have and improve our infrastructure”. —*National program manager, Lesotho

> *“We were also leveraging on machine learning, which again, in my opinion, is the future of surveillance to really predict clients who are likely to suffer unfavorable outcomes. So those who are likely to have unsuppressed viral load, those who are likely to have advanced HIV disease, those who are likely to turn positive”* —National program donor, Kenya

> *“We’re classifying people by risk using machine learning… then prioritizing limited prevention resources accordingly.”* —National program manager, Kenya

> *“We are currently doing something with WHO around some machine learning and modeling for risk. So we are able to identify people, to classify people based on their risk. And we are then able to say, in the event that you do not have enough resources for prevention, then I would focus on this group, high risk people for biomedical services”* —National program donor, Kenya

#### 3.2 Need for a tailored surveillance approach

Some respondents observed that dynamics of HIV transmissions and AIDS-related deaths are changing and will likely be geographically localized and concentrated among specific subpopulations and hence the need to devise new surveillance systems, as suggested below:

> *“All the dynamics are changing; population, social and economic… For epidemic monitoring in the future, we need systems that show us where HIV is versus where those with HIV are seeking services” —*Global Expert

> *“Prevalence has plateaued. Now we need real-time tools that capture recent infections, interruptions in treatment, and hidden populations.”* —Global Expert

> *“I think what we’re going to see is that … as treatment gets high, we’re going to have these small, small pockets of new infections that’s probably […] because that population group isn’t able to get on treatment. And so [for example] it’s the MSM in a country where MSM is highly stigmatized or illegal. And thus we will see that we need to be offering that population choices for how they prevent onward transmission. So that last mile from the, you know, 95% treatment coverage, the last 5% is going to be tricky. And I think the only way we’ll do that well is if we understand what’s needed, what choices are needed for those populations” —*Global Expert

Many respondents acknowledged that population-based surveys will unlikely be sustainable in the future because of the reduction of funding for HIV programs and declining HIV infections and deaths. Most also reported the need to continue to prioritize surveys targeting high-risk groups and sub-populations known to have poor access to health facilities and case-based surveillance for ART clients with poor outcomes, leveraging on individual level data platforms such as the EMR, recency tests and technology to efficiently digitally capture routine patient data. A few of the international and regional respondents recommended case-investigation for newly HIV-infected clients and those with advanced HIV disease to identify their uptake of and adherence to specific preventive and treatment interventions and challenges experienced with their use.

> *“[in nationwide PHIAs] we are expecting to get around 1600 [new] HIV positive clients, and we are going to shape the story for the country based on only 1600 [people]. Yet we have 1.3 million people who are on ART, whose story we have because we have the individual level data. So, for us, we arrive to the fact that if we implement our case surveillance, which we have revamped most recently, we have very rich data that not even KenPHIA can be able to tell us.”* —National policymaker, Kenya

> *“What we need in future is to have targeted PHIAs. So, recency testing combined with targeted PHIAs should be the way we handle things in the future, because we would be targeting specific geographical locations and specific subpopulations” —*National program donor, Malawi

> *“DHS in Harare just did this survey where they asked at the last sex, did you use a condom? Did you use PrEP? Were you circumcised? And tried to figure out what were the risks of that last sex act. I think that is going to be a potential for the future, is understanding what people are using for prevention and which groups of populations are using those, and then showing the importance of different choices for people, for prevention”* —Global expert

#### 3.3 Leveraging individual-level electronic data systems

Interviewees acknowledged the potential of individual-level electronic data systems to enhance surveillance, and the current efforts to expand their use in the health sector, but global experts also questioned the feasibility of scaling up and maintaining these platforms in the routine health system because of the resource requirements and ongoing government commitment required.

> *“EMRs are very hard to implement, but if they were well managed, then it would unleash a much better understanding of EVERYTHING. Ministries of Finance should be demanding this as a way to demonstrate return on investment”* —Global expert

> *“We have all the technology; we just need skilled people to do the job” —*Global expert

> *“We had thought that as countries moved to EMR, that that would be the answer. But I think we’re finding because for whatever reason, whether its people registering at multiple sites or whatever, that’s not entirely the answer. […] You know, [in Country X], they implemented this whole system of fingerprint ID for patients on ART. And they thought that that was going to be the gold standard. And it turned out to not work at all.”* —Global expert

#### 3.4 Conflicting perspectives on the importance of mortality surveillance

Several respondents expressed concern over potential inaccuracy of modelled mortality estimates and emphasized the need for renewed investments in mortality surveillance systems while a few disagreed. Proponents justified the investments as it will not only assess the impact of the treatment program and allow for development of interventions to reduce mortality but also to improve validation of mathematical models’ estimates on mortality. Opponents were concerned with the huge investments required to sustain good mortality surveillance systems and argued that some proxy indicators for mortality are already available.

> *“Mortality surveillance has not historically played much of a role [in HIV surveillance], but vital statistics are absolutely critical. But, until governments see the need for the data, that’s not going to move forward. There are opportunities for governments to push this forward, but it can’t be funded by [donors]”* —Global expert

> *“Improving mortality surveillance would help us validate the models better. ..So right now we estimate mortality, and the estimate relies on two sources. One is for people who are not on treatment, [and for those] we’re relying on mortality progression and mortality rates that came from the cohort studies, you know, in places like Rakai and other places that all took place before ART became available. So we’re relying on fitting to the survival curves from those studies for our estimates of mortality of people not on treatment. Then for mortality of people on treatment, we rely on data from the IDEA consortium …. that collects data from treatment sites around the world… and in most places, we just accept those data because we don’t have any better data to validate*” —Global expert

> *“I think it would be great if we had case surveillance and mortality surveillance, that would be very helpful. But I think we can survive without it. I think we’ve got fairly good systems in place that help us understand the epidemic and what’s happening with it, and viral load suppression, instead of waiting for deaths [to occur].”* — Global expert

#### 3.5 Streamlined HIV surveillance indicators

Most national respondents recommended that future priority indicators should include the 95-95-95 treatment cascade targets, HIV prevalence and incidence. However, many did not elaborate on the feasibility of measuring these indicators in the absence of population-based surveys. A few noted the challenges of empirically measuring HIV incidence and proposed expanding the use of the “recency HIV testing algorithm” to estimate the number of new infections and HIV incidence or using STI indicators as proxies for HIV infection. Among the selected priority surveillance indicators, most national and international respondents stressed the importance of tracking viral load suppression using the routine health facility data sources, as it is a strong predictor of HIV incidence. A few participants also emphasized the need to improve the coverage and frequency of viral load tests, quality of routine viral load data, and turnaround time of viral load results if countries are to ensure a timely public health response.

> *“As long as the percentage of people on treatment tested for viral load is high, you know, 85% or higher, we feel that’s a pretty good estimate of what’s really going on in the population, So I think viral suppression is vitally important for our estimates around the future of the epidemic. I think we feel more confident about those [viral suppression] numbers than we do about a lot of other things.”* —Global expert

> *“Access to viral load testing [is important in the future], if we can at least reduce the time [between tests]. Because, currently, the standard is a person is subjected to viral load testing once every six months. But six months seems to be quite a long time taking into account that the majority of the people who are in the last 95 are healthy people. The chances of them becoming negligent is high. So, at least reducing the time of monitoring their viral load would be something that is good. Maybe I would also suggest that we introduce CD4 monitoring as well within that group. Although we know that they are virally suppressed. But a CD4 count would measure to what extent is the HIV advancing.”* —National program implementer, Malawi

> *“As the program is also now moving towards sustainability, bringing back the STIs, which were sort of like forgotten we can still utilize our STI surveillance to be able to also identify people who are now at risk of acquiring HIV… And also, STI surveillance will also help us to zoom in, and we can be able to actually capture at an early stage, say, maybe any transmission patterns that would be happening that could also start bringing [] an increase in [HIV] incidence* —National policymaker, Kenya

Considering that a large proportion of PLHIV are now on ART, most respondents recommended close monitoring of indicators that reflect the quality, coverage and implementation challenges of the ART program such as CD4 cell counts, advanced HIV disease (AHD), loss-to-follow up, treatment adherence, ARV drug resistance, non-communicable diseases and mortality. In addition, in view of the current scale-up of various PrEP formulations, most respondents recommended close tracking of PrEP continuity instead of focusing just on the absolute number of PrEP initiations.

> *“…we should get back to CD4 testing among new diagnoses, which allows us to infer the lag time between infection and diagnosis..… that will go a long way towards what we need in the future.*” —Global expert

> *“Over time and especially in a country like ours where we have an aging epidemic, I think the incidence of non-communicable diseases are equally important. So bringing that out and including it as part of the sentinel events is important.”* —National program donor, Kenya

> *“What we’re getting from programs are reports of people newly initiated on PrEP, you know, and that’s okay. […] But that doesn’t tell us very much. So we have to rely on some special studies that have looked at continuation. How long do people stay on prep once they initiate? We’ve been trying to get recommendations for PrEP reporting changed so that we drop this crazy indicator about number of initiations and start to look at number of person months on prep somehow maybe through PrEP dispensed or whatever.”* —Global expert

> *So I think a lot of information [is needed] around progression to advanced HIV disease. I think there is something that is being called the fourth 95. I don’t know whether it’s official, but I’ve heard it somewhere in a forum [perhaps] that is mortality. So it’s that whole conversation around the people who are being lost and the ones who are dying.”*—National policymaker, Kenya

## Discussion

Our study with national, regional and international HIV stakeholders found that HIV programs in Sub-Saharan Africa have successfully used data from the routine health system, population-based surveys and mathematical modelling for HIV surveillance and M&E. We found stakeholders were concerned with the parallel systems for HIV surveillance and its lack of integration with the broad health surveillance systems.

They also noted sub-optimal quality of routine facility-based data, and unsustainability of population-based surveys, which together could negatively affect the accuracy of future estimates derived through mathematical models. They recommended increased investment in several areas including data quality improvement and adoption of digital systems, capacity building in mathematical modelling, implementation of targeted surveys focusing on high-risk populations and prioritization of specific indicators. Although the use of electronic data collection and reporting systems in the routine health systems, such as EMR and ScanForm, had expanded, they had not been fully exploited to improve surveillance.

At national and regional levels, few participants could clearly distinguish M&E activities from surveillance and often used these terms interchangeably. This finding reflects a broader challenge observed in global health, where the boundaries between routine program monitoring and disease surveillance are frequently blurred [8], a challenge exacerbated by the recommended use of the same indicators for both M&E and surveillance [9]. While surveillance’s purpose is primarily to monitor disease trends in a population, M&E activities are program-driven processes aimed at monitoring service delivery and intermediate health outcomes [10, 11], and confusion between the two purposes may contribute to fragmented systems. This lack of clarity has important implications: if M&E and surveillance are not conceptually and operationally distinguished, investments may prioritize donor reporting requirements over the development of sustainable health data systems [12], which can reduce our ability to inform epidemic control at the population level [11], and broader health system responses.

Our findings highlight both the achievements and persistent challenges of HIV surveillance and M&E systems in sub-Saharan Africa. Stakeholders acknowledged that triangulation of routine health facility data, population-based surveys, and mathematical modelling has been instrumental in tracking HIV incidence, prevalence, and programmatic outcomes, as has been documented previously [13–15]. These sources have enabled countries to monitor progress toward epidemic control and inform resource allocation. However, our participants noted the natural tension that arises from HIV M&E systems which function parallel to integrated national health information systems. This is consistent with previous evaluations noting the fragmentation of HIV-specific M&E away from integrated national health surveillance frameworks [16–18]. With the future of global funding for HIV programs still in flux, national HIV programs may need to reprioritize and pare down the indicators collected, integrating with national health information systems while ensuring they are able to keep monitoring HIV services, and collecting the minimally required indicators for surveillance purposes.

Large-scale population-based surveys, including DHS and PHIAs, provide high-quality prevalence estimates [19] but are resource-intensive and infrequent, limiting their utility for real-time program planning. Participants’ emphasis on the importance of continuing focused surveys on key and priority populations reflect global debates on the limitations of aggregated national prevalence estimates and the urgent need for granular, population-specific, electronic data to guide tailored interventions [20, 21]. Although mathematical models for surveillance such as Spectrum and Naomi have become critical for generating national and subnational HIV estimates, their accuracy depends heavily on data quality and other assumptions, leading to uncertainty in some contexts [4, 5, 15]. Also, limited awareness of key inputs of mathematical model among many local stakeholders in our study raises questions about national capacities to sustain high quality mathematical models with limited external technical support, as expected in future.

Concerns about the quality, sustainability, and accuracy of existing data sources were widely shared, and are consistent with previous findings [17, 22–25]. Routine health facility data, though increasingly integrated and digitized through platforms such as District Health Information System (DHIS2) and electronic medical records (EMR), often suffer from incomplete reporting, limited interoperability, and inadequate validation [26, 27], and an inability to adequately disaggregate health facility reports by age, sex and other important variables.

Stakeholder recommendations for improving health information systems underscore a shift toward modernization, integration and sustainability. Participants’ emphasis on the need to integrate HIV data systems with national systems is consistent with calls to reduce verticalization, enhance efficiency, and support country-led ownership of HIV data [24, 28, 29]. Calls for investment in data quality, digital tools, and artificial intelligence/machine learning (AI/ML) also reflect a growing recognition that advanced analytics can strengthen surveillance efficiency and predictive capacity [21, 30–32]. AI/ML applications, for example, may support real-time anomaly detection, hotspot prediction, and enhanced case-based surveillance [31].

Stakeholders acknowledged that despite the expansion of EMR and digital tools for conversation of paper-based records (ScanForm), their potential remains underexploited. These systems could support more integrated, patient-level, and near real-time surveillance for multiple diseases, and improved M&E for all health programs, but would require substantial investment in infrastructure, interoperability standards, and health workforce training [27–30, 33], the resources for which are now in question.

We have two weaknesses in our study: when making recommendations on surveillance system strengthening and prioritizing indicators, the respondents were not asked to comment on the feasibility and effectiveness of their recommendations. Second, purposive selection of participants for qualitative interviews means we cannot confirm whether their perspectives are representative of all stakeholders in national, regional and global contexts.

## Conclusion

Overall, our findings underscore the importance of maintaining clarity between the distinct purposes of surveillance and M&E despite utilizing the same facility-based data, and transitioning from parallel, HIV-specific surveillance toward integrated, sustainable, and technology-enabled systems. Doing so will both improve the efficiency of HIV M&E and strengthen broader health surveillance capacities, an imperative as countries confront intersecting epidemics, expanding health system demands, rapidly growing populations, and uncertainty in future donor funding. Stakeholder concerns about data quality, the cost of large-scale surveys, and modelling accuracy highlight the need for renewed investment in digital innovation, capacity building, and targeted data collection approaches. Strengthening the use of EMR and emerging technologies such as AI/ML, alongside integration with national health surveillance, will be critical to developing a more responsive, efficient, and sustainable HIV surveillance system that supports both epidemic control and wider health system strengthening.

## Data Availability

All data produced in the present study are available upon reasonable request to the authors

