## Supplementary material #1 for "Perspectives of HIV policy makers and program implementers regarding the design and utilization of HIV surveillance systems in Sub-Saharan African countries experiencing a declining HIV epidemic: a qualitative study"

**APPENDIX 2: HIV SURVEILLANCE STAKEHOLDER INTERVIEW GUIDE**

### INSTRUCTIONS FOR INTERVIEWER

Your job as an interviewer is to facilitate honest and detailed responses about what the interviewee believes about any particular response to the questions below. This is not an exam for participants agreeing to be interviewed. There are no "right" or "wrong" answers. It is permissible to ask a participant to clarify a response if you do not understand. However, do not seek unnecessary clarification, causing the nature of the original response to change substantively. While the discussion should feel natural, avoid providing too much of your own personal insight, which may lead or sway a participant to reach your own pre-determined conclusion. Your job is to motivate the participant to expand on their own ideas and allow them to reach conclusions on their own.

The questions below have been designed as to not solicit simple "yes" and "no" answers but are open-ended in such a way that participants can answer them as they see fit, given their own experience and knowledge of the question. Your role as a guide is to keep participants from straying off topic. To accomplish this, you may ask for details, stories, anecdotes, descriptions of setting, opinions, attitudes, and perceptions about responses to answers that are already on topic. Avoid repeating a question which, you feel, has already been adequately addressed. Thus, **it is not necessary to ask each and every question in the KII guide in the sequence that has been provided**.

### INTRODUCTION

Thank you very much Prof/Dr/Mr/Mrs/Ms…………………….............................. for agreeing to share information and perspectives on a study entitled *"Perspectives of HIV policy makers and programme implementers regarding the design and utilization HIV surveillance systems in Malawi and other countries with similar HIV epidemic profiles: a qualitative study"* Please help us by answering the questions that I will ask based on your experiences, expertise and opinions learned through your work.

Please indicate whether you are willing to answer the questions below, as well as the extent to which you require your participation in this process to be confidential, by answering the following questions.

- Are willing for your name and organization to be identified as a key informant in our final report?

YES NO

- Are willing for your opinions and perspectives to be accredited to you and your organization in our final report?

YES NO

- Do you understand that if you answered "NO" to both of the above questions, that we will keep all identifiable information about you anonymous?

YES NO

---

**PARTICIPANT'S BASIC DEMOGRAPHIC DATA**

Age: ___________ years

Gender: ___________ M/F, other

Highest Education Level: ___________

Occupation: ___________

Type of organization
- Public
- Local Non-governmental organization (NGO)
- International NGO
- Civil Society Organization (CSO)
- Private
- Development Partner
- Other *(please specify)*_______________________

For how long have you been working at this organization? ___________ days/weeks/months/years *(tick applicable)*

**QUESTIONS**

1. Please can you tell me a little bit about your work, and how it relates to HIV surveillance?
    - *Probe: specify his/her role eg surveillance design, data collection, data analyses, data interpretation, program decision-making)*

2. How, if at all, have you used or referenced the global or national HIV surveillance strategies in planning or implementing your work?

3. What HIV surveillance systems or studies do you know of that are conducted routinely or periodically?
    - *Probe: Global, national, subnational efforts*

4. How, if at all, do you use data from any of these studies in your work?
    - *Probe: From which studies and how do you use the data?*
    - *Probe: what specific indicators have you used to guide your work?*

5. What do you think have been the most important indicators so far in monitoring the trend of the HIV epidemic globally and nationally?
    - *Probe for each: Why was [this] important for epidemic monitoring?*
    - *Probe: Which indicators for measuring burden of HIV*
    - *Probe: which indicators for measuring coverage of HIV interventions*
    - *Probe: which indicators for measuring the impact of HIV interventions*

6. How appropriate are the existing data sources for measuring or tracking trends in the HIV epidemic?
    - *Probe: any advantages or disadvantages in terms of cost, data completeness, reliability of results, frequency of data collection, accuracy, [ability to identify small changes in a context of high ART coverage]*

7. *With the current high coverage of HIV interventions and expected low numbers of HIV infections and complications,* what do you think are the most important aspects of HIV that Malawi needs to monitor in future so as to assess progress or failure in HIV control efforts?
    - *Probe for each: Why do you feel this is important for epidemic monitoring in the future?*

8. What indicators do you think would best track these issues?
    - *Probe for each:*
        i. *What data source would be used? (ie. Routine programme data, new surveys, the same surveys previously used?).*
        ii. *Do we need new variables within existing data sources?*
        iii. *Do we need new data sources?*
